# Socioeconomic inequalities and diabetes complications: An analysis of administrative data from Hungary

**DOI:** 10.1101/2024.09.28.24314540

**Authors:** Péter Elek, Balázs Mayer, Orsolya Varga

## Abstract

**Background:** Diabetes complications are associated with increased healthcare costs and worsened patient outcomes. In this paper we analyze how individual-level demographic and territorial-level socioeconomic and healthcare variables influence the presence and severity of diabetes complications and their relationship with mortality.

**Methods:** Our study utilizes anonymized administrative data from Hungary, containing outpatient and inpatient records of all diabetes patients between 2010 to 2017. We construct settlement-year level and individual-year level panel datasets to analyze diabetes prevalence, incidence and complications, employing Poisson and logit models to explore associations between complications and the explanatory variables. The adapted Diabetes Complications Severity Index (aDCSI) is employed to quantitatively evaluate the severity of complications by aggregating individual complication scores from ICD-10 diagnosis codes.

**Results:** Diabetes prevalence and incidence are higher in settlements with above-median unemployment rates, where patients exhibit more severe complications, as shown by higher average aDCSI scores. Among socioeconomic factors, unemployment rate is particularly associated with increased aDCSI scores, while better healthcare access is associated with lower aDCSI scores in unadjusted but with higher scores in adjusted models. The presence and severity of complications, especially renal, cardio-vascular and peripheral vascular ones, substantially increase five-year inpatient mortality. Most of the mortality difference by settlement-level unemployment rate disappears when complications are accounted for.

**Conclusions:** Territorial-level socioeconomic disparities, especially elevated unemployment rates, significantly exacerbate the severity of diabetes complications, which subsequently heightens the mortality risk among diabetic patients. Enhancing health-care access in lower socioeconomic regions may alleviate these disparities and lessen the impact of complications.

## Introduction

The IDF Diabetes Atlas 2021 projects that the prevalence of diabetes in the European Region (currently at 9.2%) will increase by 13% by 2045 [1]. The elevated risk of mortality and morbidity in patients with diabetes is well documented and has been the subject of numerous studies [2]. Hence, diabetes represents a significant focus in public health, not only in consideration of the disease itself, but also in light of its associated comorbidities and complications. Diabetes comorbidities are additional health conditions that coexist with the disease but are not directly caused by it such as hypertension or hyperlipidaemia [3]. In contrast, complications are direct health issues arising from the long-term effects of uncontrolled blood sugar levels during the course of the disease [4]. These include renal, ophthalmology, cardiovascular, cerebrovascular, peripheral vascular, neurological and metabolic complications, as summarized by the adapted Diabetes Complications Severity Index (aDCSI) [5]. Considerable effort is being invested in research on diabetes complications, with a particular focus on their economic implications [6] and development of more effective treatment strategies and improved patient outcomes [7].

There are large disparities in the occurrence of complications. The existing literature provides evidence for the association between socioeconomic status (SES), neighborhood and physical environment, food environment, healthcare, and social context with diabetes-related health outcomes [8–10]. Studies have shown that both individual and settlement-based low SES are associated with a heightened likelihood of developing diabetes complications [11–15]. In particular, settlement-based SES indicators are of significant relevance, as they can furnish a direct input for the formulation of health policy interventions [16]. A promising approach to reducing health inequalities is to target interventions at unemployment, which is a key indicator of socio-economic status (SES) and is associated with poor health [17].

In this paper we analyse the individual- and settlement-level determinants of diabetes complications in Hungary, a European Union member state with about 10 million inhabitants. Although a number of studies on particular diabetes complications have been conducted in the country [18–23], these studies devote limited attention to the relationship between SES determinants and none of them address the issue of unemployment. Our study employs a comprehensive individual-level administrative panel dataset comprising outpatient and inpatient records for all diabetes patients in Hungary over the period 2010– 2017. We provide a systematic description of the prevalence and severity of complications in the aDCSI framework and explore their association with individual-level demographic and settlement-level socioeconomic variables with a special focus on unemployment. Finally, we investigate how settlement-level variables influence the relationship between complications and mortality.

## Methods

### Institutional setting

The Hungarian state health insurance scheme provides patients with access to a comprehensive range of services, encompassing primary care, specialist outpatient care, inpatient care, and pharmaceuticals [24]. Apart from pharmaceuticals that require co-payments, these services are free of charge at the point of use. However, Hungary’s public healthcare spending is low compared to other EU countries, leading to substantial out-of-pocket expenses for private healthcare [25].

The care of patients with type 2 diabetes mellitus (T2DM) is primarily the responsibility of general practitioners (GPs), who must navigate a number of challenges, including limited time for patient care and the typical presence of only one auxiliary health worker. Furthermore, approximately 5 percent of primary care practices were vacant during our examined period, particularly in rural areas, which constrains access to primary care [26].

Several specialists are involved in diabetes care, including diabetologists for recommending prescription of drugs other than metformin and sulfonylureas, and nephrologists, ophthalmologists, cardiologists, neurologists, angiologists for managing complications. At this phase, the provision of care for individuals with diabetes is appropriately concentrated in specialist outpatient facilities [27], which are generally located in Budapest, the 19 county seats and the district seat towns (of which there are 174 outside the capital). Nevertheless, some rural districts still lack such facilities, hence patients residing there should travel to a neighbouring district seat or to a county seat to access specialist care [28]. Inpatient care is generally organized on the county level, although there exist hospitals outside the county seats as well.

### Data

We use an anonymized administrative dataset that contains detailed outpatient and inpatient records of all people in Hungary who consumed an antidiabetic medication (insulin or oral antidiabetics, ATC [Anatomical Therapeutic Chemical] A10 category) or attended outpatient or inpatient services with a diabetes diagnosis code (ICD-10 [Tenth Revision of the International Classification of Diseases] E10-E14) at least once between 2010–2017. (The dataset was available for research purposes on a restricted server at the Databank of HUN-REN Centre for Economic and Regional Studies under an agreement with the National Healthcare Services Centre.) The analysis excludes patients who were recorded with a diabetes diagnosis on at most two occasions during the specified period, but still includes patients who died in hospital within the year of their initial diagnosis. (Only in-hospital deaths are observed in the dataset.) Furthermore, women diagnosed with gestational diabetes or polycystic ovary syndrome at least once during the examined period are excluded. Only patients aged 18 years or above are included. These restrictions closely follow those of other papers using Hungarian administrative data on diabetes, such as [29].

Then we define diabetes patients for a given year as those who received antidiabetic medication, or attended outpatient or inpatient care with a diabetes diagnosis in that year. New diabetes cases are defined as those transitioning into diabetes after not being a diabetic patient for the preceding 2 years.

We evaluate the presence and overall severity of complications with the adapted Diabetes Complications Severity Index (aDCSI). First, each ICD-10 diagnosis code of Supplementary file Table S1 (adapted from [30]) is assigned a score of 1 (non-severe) or 2 (severe) if it appears in the person’s outpatient or inpatient records in the current or in the previous year. (Two-year time windows were selected for analysis because, even in the presence of chronic complications, not all patients appear in the healthcare system each year with the given diagnosis code.) Afterwards, for each of the seven examined organ systems, the largest score is retained and the aDCSI is calculated as the sum of individual complication scores. (For neurological complications, no diagnosis code is classified as severe, so the aDCSI takes values between 0 and 13. All metabolic complications are assigned a score of 2.) The validity of the aDCSI as a reliable metric for predicting hospitalization rates, mortality outcomes and healthcare costs was affirmed in various countries and healthcare settings [30–33].

In the analyses we use individual-level demographic data (gender and age) and territorial-level data as explanatory variables. The latter include settlement-level socioeconomic indicators (settlement type, unemployment rate, proportion of tertiary graduates, per capita taxable income) and various healthcare supply indicators. We measure access to primary care with the ratio of filled primary care practices (PCPs) at the settlement level, and access to specialist outpatient care and inpatient care with per capita number of outpatient hours and, respectively, per capita number of hospital beds at the district level. These territorial-level variables come from the municipal statistical system of the Central Statistical Office or from the National Health Insurance Fund Administration.

It is notable that all settlement-level indicators are available for the 23 districts of Budapest. Consequently, they are treated as settlements in this analysis. Furthermore, the smallest settlements in the country are merged into larger neighbouring settlements. Hence, we end up with 2,057 settlements, with an average population of 4,750 people.

### Statistical analysis

Based on the data above, we construct an individual-year level panel dataset of diabetic patients and a settlement-year level panel dataset of diabetes prevalence and incidence (using the population of the settlements as the denominator).

After showing descriptive statistics, first we use the individual-year level panel data to explore the association between complications and the explanatory variables. For examining the prevalent population, we use data for year 2016. When the dependent variable is the binary indicator of the presence of a specific complication, we fit logit models, while for the analysis of the aDCSI we estimate Poisson models. Explanatory variables are individual-as well as settlement-level variables, and we cluster standard errors on the settlement level. In such settings, regression modelling using clustered standard errors is a viable alternative to hierarchical modelling because it makes a smaller number of assumptions [34, 35]. Still, for robustness check we fit to aDCSI a multilevel Poisson model with settlement-level random effects.

Second, we examine the evolution of complications around the time of diabetes diagnosis. After plotting the average aDCSI and the prevalence of complications as a function of the relative time since diagnosis, we fit a Poisson model of aDCSI on settlement- and individual-level variables using the newly diagnosed population.

Finally, we examine with logit models how settlement- and individual-level variables influence inpatient mortality within five years, and how the relationships are moderated by including the presence and severity of complications measured at the beginning of the time period.

## Results

### Descriptive results

Table 1 presents descriptive statistics split by whether the settlement’s unemployment rate, a general indicator of settlement-level SES, is below or above median. Among the 18+ years old population, prevalence of diabetes stands at 7.2%, incidence at 0.65%, the average age of diabetic patients is 65.7 years, the proportion of males is 47%, average aDCSI is 1.59 (37% of patients have zero, 38% have 1 or 2 and 25% have a larger diagnosed aDCSI score) and their 5-year inpatient mortality is 14%.

**Table 1:** Descriptive statistics by settlement-level unemployment rate.

|  | Total | S.D. | Unemployment rate |  |
| --- | --- | --- | --- | --- |
|  |  |  | below med. | above med. |
| Prevalence of diabetes | 7.15% | 1.55% | 6.90% | 7.41% |
| Incidence of diabetes | 0.65% | 0.21% | 0.64% | 0.66% |
| aDCSI |  |  |  |  |
| aDCSI score | 1.59 | 1.75 | 1.49 | 1.68 |
| aDCSI score = 0 | 36.6% |  | 39.1% | 34.2% |
| aDCSI score = 1 | 21.1% |  | 21.2% | 21.1% |
| aDCSI score = 2 | 17.2% |  | 16.6% | 17.7% |
| aDCSI score = 3 | 10.2% |  | 9.5% | 10.9% |
| aDCSI score = 4 | 7.4% |  | 6.9% | 7.9% |
| aDCSI score = 5 | 3.8% |  | 3.4% | 4.2% |
| aDCSI score = 6+ | 3.6% |  | 3.3% | 3.9% |
| Prevalence of complications |  |  |  |  |
| Ocular (any) | 23.8% |  | 21.7% | 25.7% |
| Ocular (only 1) | 22.4% |  | 20.4% | 24.4% |
| Ocular (2) | 1.3% |  | 1.3% | 1.4% |
| Renal (any) | 10.3% |  | 9.7% | 10.9% |
| Renal (only 1) | 1.6% |  | 1.3% | 1.8% |
| Renal (2) | 8.7% |  | 8.3% | 9.1% |
| Cardiovascular (any) | 41.4% |  | 38.3% | 44.2% |
| Cardiovascular (only 1) | 25.2% |  | 22.9% | 27.4% |
| Cardiovascular (2) | 16.1% |  | 15.4% | 16.9% |
| Cerebrovascular (any) | 14.0% |  | 13.1% | 14.9% |
| Cerebrovascular (only 1) | 4.2% |  | 4.0% | 4.4% |
| Cerebrovascular (2) | 9.8% |  | 9.1% | 10.4% |
| Peripheral vascular (any) | 8.1% |  | 7.6% | 8.6% |
| Peripheral vascular (only 1) | 4.0% |  | 3.7% | 4.3% |
| Peripheral vascular (2) | 4.1% |  | 4.0% | 4.3% |
| Neurological (1) | 18.9% |  | 18.3% | 19.4% |
| Metabolic (2) | 1.2% |  | 1.2% | 1.2% |
| Demographic variables |  |  |  |  |
| Age | 65.7 | 12.2 | 66.2 | 65.2 |
| Gender: male | 47.2% |  | 48.1% | 46.4% |
| Age at diagnosis | 60.3 | 13.0 | 60.7 | 59.9 |
| Age at hospital death | 75.6 | 10.2 | 76.1 | 75.1 |
| Inpatient mortality within 5 year (2011) | 14.0% |  | 14.1% | 13.9% |
| Settlement-level SES variables |  |  |  |  |
| Unemployment rate | 6.8% | 4.2% | 3.8% | 9.6% |
| Ratio of graduates | 18.3% | 10.8% | 24.4% | 12.5% |
| Log income per cap. | 13.53 | 0.314 | 13.74 | 13.33 |
| Settlement type |  |  |  |  |
| Budapest | 16.8% |  | 34.2% | 0.0% |
| County seat | 17.1% |  | 20.0% | 14.4% |
| Other | 66.1% |  | 45.8% | 85.6% |
| Healthcare supply variables |  |  |  |  |
| Ratio of filled-in primary care practices | 95.3% | 14.9% | 97.8% | 92.8% |
| Outpatient hours per cap. | 1.92 | 1.24 | 2.36 | 1.49 |
| Hospital beds per 1000 cap. | 6.98 | 5.90 | 8.06 | 5.94 |
| Number of individuals | 659,340 |  | 323,138 | 336,202 |

Descriptive statistics in total and by settlement-level unemployment rate (below or above median). Standard deviation (S.D.) is shown for non-binary variables. Population: diabetic population in 2016, unless indicated otherwise.

Among the particular complications, cardiovascular diagnoses, including AMI, heart failure, ischemic heart disease and atrial fibrillation, are the most common (41% for any and 16% for severe complications). The prevalence of ocular complications (mainly retinopathy and its consequences), cerebrovascular complications (mainly stroke and transient ischemic attack) and neurological complications (mainly diabetic neuropathy) is 14–24% each. Less common complications include renal and peripheral vascular (8–9%) and metabolic (1%) manifestations. Supplementary file Table S2 shows that the individual complications are relatively mildly correlated: only the correlation of cardiovascular manifestations with renal and cerebrovascular complications is noteworthy (0.22–0.23).

According to Table 1, settlements with above-median compared to below-median unemployment rates have larger prevalence and incidence of diabetes, a larger average aDCSI score and larger probabilities of individual complications. At the same time, the average age of patients is lower there by 1.0 year because both the age of diagnosis and the age of death are lower. As a combined consequence of the above two findings, raw inpatient mortality is similar in the two groups.

Finally, the table shows that healthcare supply and thus access to care is negatively related to unemployment rate. The ratio of filled PCPs, the district-level outpatient hours per capita and hospital beds per capita are all much lower in settlements with unemployment rate above the median. These settlements mostly lie outside Budapest and the county seats.

### Regression results

#### Diabetes complications

Table 2 shows risk ratios from Poisson regressions of aDCSI on territorial- and individual-level variables. First, we interpret models for the prevalent population of 2016. Column (1) displays results from the unadjusted regressions, in which only a set of territorial-level variables is controlled for at a time. If unemployment rate is 1% point larger then age- and gender-controlled aDCSI is 2.1% larger (95% confidence interval [CI] 1.8%–2.1%), if the ratio of graduates is 1% point larger then aDCSI is 0.4% smaller (95% CI 0.2%–0.6%), and if income is 10% larger then aDCSI is 2.4% smaller (95% CI 1.8%–2.6%). In comparison to other parts of the country, patients in Budapest have a 8.5% smaller aDCSI (95% CI 4.3%– 12.5%). Regarding healthcare supply variables, the ratio of filled-in PCPs in the settlements have a negative association with aDCSI, e.g. a settlement with all its PCPs filled has a 9.5% lower aDCSI than a settlement with all its PCPs unfilled (95% CI 5.0%–13.8%). The associations with the other healthcare supply variables are not significant. These results suggest that in settlement with worse socioeconomic conditions and lower access to care, aDCSI is higher.

**Table 2:** Risk ratios (with 95% confidence intervals) from Poisson regressions of aDCSI on territorial- and individual-level variables.

| Risk ratios | (1)<br>unadjusted | (2)<br>adjusted<br>diabetic population | (3)<br>adjusted | (4)<br>adjusted<br>1yr after diagnosis |
| --- | --- | --- | --- | --- |
| **Dependent variable: aDCSI** |  |  |  |  |
| **Settlement-level SES variables** |  |  |  |  |
| (A) Unemployment rate (%) | 1.021<br>(1.018 - 1.023) | 1.021<br>(1.017 - 1.025) | 1.024<br>(1.013 - 1.036) | 1.024<br>(1.020 - 1.028) |
| (B) Ratio of graduates (%) | 0.996<br>(0.994 - 0.998) | 1.002<br>(1.000 - 1.004) | 1.003<br>(0.999 - 1.006) | 1.002<br>(0.999 - 1.004) |
| (C) Log income per capita $\times 10$ | 0.976<br>(0.974 - 0.982) | 0.994<br>(0.986 - 1.002) | 0.983<br>(0.968 - 0.999) | 0.998<br>(0.990 - 1.006) |
| (D) Settlement type (ref = other) |  |  |  |  |
| Budapest | 0.915<br>(0.875 - 0.957) | 0.906<br>(0.840 - 0.977) | 0.902<br>(0.820 - 0.992) | 0.894<br>(0.818 - 0.976) |
| County seat | 0.979<br>(0.897 - 1.069) | 0.955<br>(0.886 - 1.029) | 0.944<br>(0.871 - 1.024) | 0.923<br>(0.850 - 1.002) |
| (E) Healthcare supply variables |  |  |  |  |
| Ratio of filled-in primary care practices (%) | 0.905<br>(0.862 - 0.950) | 1.029<br>(0.987 - 1.073) | 1.183<br>(1.023 - 1.367) | 1.025<br>(0.975 - 1.077) |
| Outpatient hours per capita | 0.989<br>(0.960 - 1.019) | 1.052<br>(1.020 - 1.085) | 1.075<br>(1.031 - 1.122) | 1.055<br>(1.015 - 1.096) |
| Hospital beds per 1000 capita | 1.002<br>(0.997 - 1.007) | 0.997<br>(0.993 - 1.002) | 0.996<br>(0.989 - 1.002) | 0.998<br>(0.993 - 1.003) |
| Gender (ref = female) |  |  |  |  |
| Gender: male | yes | 1.125<br>(1.116 - 1.133) | 1.126<br>(1.118 - 1.134) | 1.158<br>(1.145 - 1.171) |
| Age group (ref = 60-69) |  |  |  |  |
| Age group: 18-49 | yes | 0.432<br>(0.426 - 0.439) | 0.433<br>(0.427 - 0.440) | 0.390<br>(0.382 - 0.398) |
| Age group: 50-59 | yes | 0.798<br>(0.790 - 0.807) | 0.798<br>(0.790 - 0.807) | 0.761<br>(0.748 - 0.773) |
| Age group: 70-79 | yes | 1.316<br>(1.304 - 1.328) | 1.317<br>(1.305 - 1.329) | 1.424<br>(1.403 - 1.446) |
| Age group: 80+ | yes | 1.533<br>(1.507 - 1.559) | 1.533<br>(1.507 - 1.559) | 1.914<br>(1.870 - 1.959) |
| Variance of random effect |  |  | 1.038<br>(1.028 - 1.049) |  |
| Average aDCSI | 1.59 | 1.59 | 1.59 | 1.16 |
| Number of individuals | 659,340 | 659,156 | 659,156 | 278,155 |
| Number of settlements |  |  | 2,057 |  |

95% confidence intervals are in parentheses.
**Models.** All columns except (3): cross-sectional models with standard errors clustered at the settlement level. Column (3): multilevel model with settlements as random effects.
**Population.** Columns (1)-(3): diabetic population in 2016. Column (4): population one year after a new diabetes diagnosis in years 2011-2017.
**Controls.** Gender and age group are always controlled for. Column (1): unadjusted models where a set of territorial-level variables are controlled for in each case (A, B, C, D, E). Columns (2)–(4): adjusted models where all territorial-level variables are controlled for simultaneously. Column (4): calendar years are additional controls.

According to Supplementary file Table S3, the settlement-level SES variables demonstrate a high degree of correlation. Therefore, it seems prudent to investigate which socioeconomic variables have a direct influence on the number and severity of complications. Columns (2)–(3) of Table 2 show the results from the adjusted Poisson models in which the territorial-level variables are included simultaneously. (A model with standard errors clustered at the settlement level is used in column (2), while a hierarchical model with settlements as random effects is used in column (3) for robustness check. The results of the two models do not differ substantially.)

After controlling for other factors, males have 12.5% larger aDCSI than females (95% CI 11.6%– 13.3%), and there is a steep age gradient (e.g. 80+ years old patients have 53% larger aDCSI than the 60-69 years old patients (95% CI 50.7%–55.9%)). A 1% point larger unemployment rate is associated with a 2.1% larger aDCSI (95% CI 1.7%–2.5%), and the coefficients of the ratio of graduates and log income per capita are not statistically significant at the 5% level in column (2).

After controlling for socioeconomic conditions, outpatient hours per capita (and in one specification the ratio of filled PCPs) is positively associated with aDCSI, which may be a consequence of the under-diagnosis of these conditions in the absence of appropriate access to healthcare. Finally, the negative coefficient of Budapest remains essentially unchanged in the adjusted models as well.

Table 3 displays odds ratios (ORs) from logit models for the presence of complications by organ system. The largest ORs of male gender (1.59 and 1.29) are observed for peripheral vascular and cardiovascular complications, while gender differences are small for the other complications. Regarding age groups, the steepest positive age gradient (OR above 1.8 for the 80+ compared to the 60-69 years old population) is estimated for renal, cardiovascular and cerebrovascular complications, while a clear negative gradient of age is observed for metabolic complications (affecting mostly type 1 diabetes patients). With regard to the SES variables, the ORs of a 1%point change of the unemployment rate are statistically significant and are in a relatively narrow range (1.02–1.05), apart from the case of metabolic complications. The adjusted ORs for the remaining SES variables are less clear-cut.

**Table 3:** Adjusted odds ratios (with 95% confidence intervals) from logistic regressions of the presence of complications on territorial- and individual-level variables.

| Odds ratios | Ocular | Renal | Cardio | Cerebro | Periph | Neuro | Metab |
| --- | --- | --- | --- | --- | --- | --- | --- |
| Settlement-level SES variables |  |  |  |  |  |  |  |
| Unemployment rate (%) | 1.048<br>(1.029 - 1.068) | 1.033<br>(1.021 - 1.046) | 1.034<br>(1.024 - 1.045) | 1.025<br>(1.014 - 1.036) | 1.031<br>(1.018 - 1.044) | 1.024<br>(1.015 - 1.034) | 1.006<br>(0.973 - 1.041) |
| Ratio of graduates (%) | 1.005<br>(0.992 - 1.018) | 1.002<br>(0.996 - 1.007) | 1.008<br>(1.004 - 1.012) | 1.002<br>(0.996 - 1.007) | 0.995<br>(0.989 - 1.000) | 1.005<br>(1.000 - 1.009) | 1.000<br>(0.984 - 1.017) |
| Log income per cap. $\times 10$ | 0.996<br>(0.956 - 1.038) | 1.018<br>(0.994 - 1.043) | 0.966<br>(0.945 - 0.987) | 0.991<br>(0.971 - 1.012) | 1.016<br>(0.989 - 1.043) | 1.001<br>(0.983 - 1.019) | 0.987<br>(0.930 - 1.048) |
| Settlement type (ref = other) |  |  |  |  |  |  |  |
| Budapest | 1.205<br>(0.787 - 1.847) | 0.707<br>(0.551 - 0.907) | 0.849<br>(0.741 - 0.973) | 0.829<br>(0.691 - 0.993) | 0.923<br>(0.713 - 1.195) | 0.824<br>(0.663 - 1.025) | 0.895<br>(0.495 - 1.618) |
| County seat | 0.987<br>(0.685 - 1.422) | 0.945<br>(0.808 - 1.106) | 0.952<br>(0.832 - 1.090) | 0.877<br>(0.752 - 1.023) | 0.922<br>(0.706 - 1.203) | 0.890<br>(0.766 - 1.033) | 1.313<br>(0.717 - 2.405) |
| Healthcare supply variables |  |  |  |  |  |  |  |
| Filled-in PCP ratio | 1.335<br>(1.116 - 1.598) | 0.913<br>(0.825 - 1.011) | 1.055<br>(0.960 - 1.159) | 1.025<br>(0.929 - 1.130) | 1.026<br>(0.878 - 1.198) | 0.968<br>(0.865 - 1.084) | 0.985<br>(0.691 - 1.405) |
| Outpatient hours per cap. | 0.975<br>(0.812 - 1.171) | 1.127<br>(1.001 - 1.269) | 1.096<br>(1.039 - 1.156) | 1.033<br>(0.960 - 1.111) | 1.064<br>(0.934 - 1.212) | 1.126<br>(1.003 - 1.265) | 1.208<br>(0.928 - 1.571) |
| Hosp. beds per 1000 cap. | 1.011<br>(0.984 - 1.038) | 0.992<br>(0.978 - 1.007) | 0.991<br>(0.984 - 0.997) | 1.005<br>(0.994 - 1.016) | 0.997<br>(0.978 - 1.017) | 0.988<br>(0.973 - 1.004) | 0.965<br>(0.925 - 1.007) |
| Gender (ref = female) |  |  |  |  |  |  |  |
| Gender: male | 0.991<br>(0.973 - 1.010) | 1.074<br>(1.051 - 1.098) | 1.287<br>(1.265 - 1.309) | 0.993<br>(0.972 - 1.015) | 1.586<br>(1.549 - 1.625) | 0.944<br>(0.929 - 0.960) | 1.026<br>(0.981 - 1.074) |
| Age group (ref = 60-69 years) |  |  |  |  |  |  |  |
| Age group: 18-49 | 0.550<br>(0.531 - 0.571) | 0.490<br>(0.467 - 0.513) | 0.197<br>(0.191 - 0.203) | 0.224<br>(0.213 - 0.235) | 0.391<br>(0.374 - 0.410) | 0.491<br>(0.477 - 0.506) | 2.437<br>(2.213 - 2.683) |
| Age group: 50-59 | 0.849<br>(0.832 - 0.867) | 0.673<br>(0.653 - 0.694) | 0.625<br>(0.614 - 0.636) | 0.702<br>(0.684 - 0.721) | 0.853<br>(0.828 - 0.879) | 0.971<br>(0.951 - 0.992) | 1.228<br>(1.147 - 1.315) |
| Age group: 70-79 | 1.132<br>(1.106 - 1.158) | 1.912<br>(1.859 - 1.967) | 1.636<br>(1.612 - 1.660) | 1.497<br>(1.466 - 1.529) | 1.133<br>(1.106 - 1.161) | 1.085<br>(1.062 - 1.108) | 0.962<br>(0.889 - 1.042) |
| Age group: 80+ | 0.922<br>(0.875 - 0.971) | 3.163<br>(3.045 - 3.285) | 2.317<br>(2.249 - 2.387) | 1.831<br>(1.777 - 1.887) | 1.163<br>(1.122 - 1.206) | 0.917<br>(0.884 - 0.951) | 0.784<br>(0.719 - 0.856) |
| Probabilities | 23.8% | 10.3% | 41.4% | 14.0% | 8.1% | 18.9% | 1.2% |
| Number of individuals | 659,156 | 659,156 | 659,156 | 659,156 | 659,156 | 659,156 | 659,156 |

95% confidence intervals calculated from settlement-level clustered standard errors are in parentheses.
Population: diabetic patients in 2016.

#### Complications around the time of diabetes diagnosis

Given the longitudinal nature of the data, the complications can also be examined on the newly diagnosed population. Column (4) of Table 2 presents risk ratios derived from Poisson regressions of aDCSI on territorial- and individual-level variables one year after the diagnosis. As might be expected, the mean aDCSI is slightly smaller in this population than in the prevalent population (1.16 vs. 1.59). The gender and age gradient are more pronounced around the time of diagnosis than for the prevalent population (columns (2)–(3)). The effect of the territorial-level variables is not significantly different. For example, the risk ratio for the unemployment rate (1.024, 95% CI 1.020–1.028) is not notably disparate from the parameter measured on the prevalent population.

Supplementary file Figure S1 shows the evolution of aDCSI and the individual complication probabilities by relative year since the diagnosis of diabetes. The average aDCSI score of 0.6 before the diagnosis jumps to around 1.2 around the time of diagnosis, suggesting that some of the conditions had already been detected earlier. The score decreases slightly after two years because the detection of some complications coincides with the diagnosis, and later a small proportion of complications may be missing from the administrative records due to not attending healthcare facilities, but the score remains roughly constant in relative years 2 to 4.

Looking at individual complication probabilities, renal, peripheral vascular, neurological and (not surprisingly) metabolic complications often get detected around the time of diagnosis – their observed prevalence jumps more than twofold then. Meanwhile, cerebrovascular complications do not become drastically more frequent around the time of diagnosis.

#### Inpatient mortality

Table 4 displays the socioeconomic, demographic and complication-related determinants of five-year inpatient mortality. Beyond the well-known effects of age group and gender, the value of aDCSI has a marked impact on mortality within 5 years. According to column (2), compared to a score of 0, OR=1.053 (95% CI 1.028–1.079) for a score of 1 but OR=1.487 (95% CI 1.449–1.526) for a score of 2, and OR=1.234 (95% CI 1.223–1.245) for each additional point in the score. Nevertheless, these average effects of aDCSI hide a marked variation across individual complication categories. Column (3) shows that renal, cardiovascular and peripheral vascular complications have the strongest association with mortality, with OR between 1.1–1.3 for non-severe (aDCSI=1) and 1.8–2.1 for severe (aDCSI=2) manifestations. Ocular, cerebrovascular and neurological complications have smaller effects (OR close to 1 or even negative for non-severe and 1.4–1.5 for severe manifestations). Metabolic complications (that occur infrequently) do not have a significant impact on mortality.

**Table 4:** Adjusted odds ratios (with 95% confidence intervals) from logistic regressions of inpatient mortality within five years on territorial- and individual-level variables.

| Odds ratios | (1) | (2) | (3) |
| --- | --- | --- | --- |
| Dep. var.: 5-year inpatient mortality |  |  |  |
| Unemployment rate (%) | 1.011<br>(1.008 - 1.014) | 1.003<br>(1.000 - 1.007) | 1.004<br>(1.000 - 1.007) |
| aDCSI (ref = 0) |  |  |  |
| aDCSI=1 |  | 1.053<br>(1.028 - 1.079) |  |
| aDCSI=2 |  | 1.487<br>(1.449 - 1.526) |  |
| aDCSI, additional point |  | 1.234<br>(1.223 - 1.245) |  |
| Diabetes complications |  |  |  |
| Ocular (only 1) |  |  | 0.959<br>(0.928 - 0.992) |
| Renal (only 1) |  |  | 1.285<br>(1.215 - 1.359) |
| Cardiovascular (only 1) |  |  | 1.169<br>(1.144 - 1.194) |
| Cerebrovascular (only 1) |  |  | 0.876<br>(0.840 - 0.913) |
| Peripheral vascular (only 1) |  |  | 1.237<br>(1.178 - 1.299) |
| Neurological (only 1) |  |  | 1.045<br>(1.020 - 1.069) |
| Ocular (2) |  |  | 1.497<br>(1.415 - 1.583) |
| Renal (2) |  |  | 1.793<br>(1.717 - 1.872) |
| Cardiovascular (2) |  |  | 1.971<br>(1.928 - 2.016) |
| Cerebrovascular (2) |  |  | 1.415<br>(1.380 - 1.450) |
| Peripheral vascular (2) |  |  | 2.109<br>(2.042 - 2.179) |
| Metabolic (2) |  |  | 1.081<br>(0.964 - 1.214) |
| Gender (ref = female) |  |  |  |
| Gender: male | 1.436<br>(1.413 - 1.460) | 1.375<br>(1.352 - 1.399) | 1.338<br>(1.315 - 1.360) |
| Age group (ref = 60-69) |  |  |  |
| Age group: 18-49 | 0.209<br>(0.198 - 0.220) | 0.245<br>(0.233 - 0.259) | 0.249<br>(0.236 - 0.262) |
| Age group: 50-59 | 0.557<br>(0.543 - 0.572) | 0.578<br>(0.564 - 0.593) | 0.590<br>(0.575 - 0.606) |
| Age group: 70-79 | 2.336<br>(2.287 - 2.387) | 2.150<br>(2.103 - 2.198) | 2.112<br>(2.067 - 2.159) |
| Age group: 80+ | 5.501<br>(5.356 - 5.651) | 4.959<br>(4.831 - 5.091) | 4.706<br>(4.584 - 4.832) |
| Number of individuals | 644,880 | 644,880 | 644,880 |

95% confidence intervals calculated from settlement-level clustered standard errors are in parentheses. Population: diabetic patients in 2011.

Among the territorial explanatory variables, for ease of interpretation we only use the unemployment rate in these regressions. If complications are not controlled for, a 1%point larger unemployment rate is associated with a 1.1% larger odds of five-year inpatient mortality (OR=1.011, 95% CI 1.008–1.014). However, if complications are controlled for, it increases the odds by only 0.3–0.4% (95% CI for OR: 1.000–1.007). This suggests that most of the mortality differences of diabetes patients by settlement-level unemployment rate is attributable to the fact that patients in lower-SES settlements are affected more severely by diabetes complications.

## Discussion

This is the first study to utilise complete administrative data, aligning with the aDCSI definitions, to investigate complications in both prevalent and incident diabetic populations, as well as mortality in Hungary. Additionally, it incorporates settlement-level variables, offering a nuanced understanding of the complex relationships between complications and their determinants.

Among diabetic patients, the average value of aDCSI is 1.6. Cardiovascular complications are the most prevalent (41%), while ocular, cerebrovascular, and neurological complications also commonly occur (14-24%), underscoring the significant morbidity of diabetes [36]. Patients in lower-SES settlements that are characterized by higher unemployment rates, fewer degree-holders and lower per capita income have substantially higher average scores on the aDCSI, indicating greater diagnosed complication burden, likely due to delayed diagnosis, suboptimal management, and restricted healthcare access [37]. Indeed, disadvantaged areas have fewer primary care physicians, outpatient hours, and hospital beds per capita.

The results of the Poisson and logit models demonstrate the independent influence of socioeconomic and demographic factors on the severity of diabetes complications in the studied population. In particular, a one standard deviation (4.2%point) increase in the unemployment rate is associated with a 9.1% increase in the adjusted aDCSI, which demonstrates the substantial influence of settlement-level socioeconomic circumstances on health outcomes. The aDCSI is lower in Budapest, which is partly attributable to enhanced accessibility to healthcare resources. The adjusted models indicate that males have a 13% higher aDCSI than females, and individuals aged 80 and above experience a 53% increase in aDCSI compared to those aged 60-69. These findings underscore the necessity to tackle socioeconomic disparities and enhance healthcare accessibility with a view to mitigating the adverse impact of diabetes complications [11, 12].

The considerable rise in aDCSI around diagnosis emphasises the necessity for prompt detection and management of complications to enhance patient outcomes. However, SES disparities persist over time, as evidenced by the comparable effects of settlement-level variables at diagnosis versus in the prevalent population. Longitudinal analysis offers valuable insights into the complex interplay between disease progression and healthcare access in shaping diabetes complication trajectories.

Finally, our mortality regressions show that the higher complication burden of lower-SES settlements largely explain the higher mortality rates observed there, highlighting the need for targeted healthcare interventions in disadvantaged areas [5, 14]. The substantially different contribution of the organ-specific complication scores to mortality suggest that at least for predicting mortality in the Hungarian population, renal, cardiovascular and peripheral vascular complications could be assigned more weight than the others, and that severe complications could be assigned more than twice the weight of non-severe complications. These findings underscore the importance of healthcare providers prioritising the most impactful complications when assessing patient risk and developing treatment plans, particularly in populations with high rates of diabetes and related complications [5, 38].

Our systematic analysis of diabetes complications using the aDCSI score and a comprehensive set of nationally representative longitudinal data makes a significant contribution to the literature. However, the study has certain limitations. First, due to the possibility of under-diagnosis, administrative data may not capture all complications present. Second, we observe only inpatient mortality and not total mortality, however, approximately 65% of individuals die in hospital in Hungary. Finally, information on individual-level SES would be useful to examine the causal effect of SES on the prevalence and severity of complications in a longitudinal setting.

## Statements

### Funding

The research was supported by National Research, Development and Innovation Office grant OTKA FK 134573.

### Conflicts of interest

none declared.

### Data availability

The anonymized data used in this article were provided by the National Healthcare Services Centre (ÁEEK) under an agreement with HUN-REN Centre for Economic and Regional Studies (KRTK), and were processed on a restricted server at the Databank of HUN-REN KRTK. The data cannot be shared with outside parties. Other, territorial-level data used in this article may be shared on request with permission of HUN-REN KRTK.

### Ethical approval

The use of the anonymized dataset for research purposes was approved by the Hungarian Medical Research Council (ETT TUKEB, 39258/2019/EKU).

### Author Contributions

PE and BM were the principal contributors to the data cleansing and statistical analysis. OV played an integral role in steering the study and defining the final submission. Collectively, PE, BM, and OV collaborated on the interpretation of the data and wrote the manuscript. All authors provided invaluable feedback on the manuscript, ensuring its intellectual rigour and quality.

## Key points

- Nationwide individual-level administrative panel data from Hungary are used to analyze diabetes complications.
- The adapted Diabetes Complications Severity Index (aDCSI) is employed.
- Among settlement-level socioeconomic variables, unemployment rate is particularly associated with aDCSI.
- Most of the mortality difference by settlement-level unemployment rate disappears when complications are accounted for.

## Notes

### Competing Interest Statement

The authors have declared no competing interest.

### Author Declarations

Ethics committee of the Hungarian Medical Research Council gave ethical approval for this work. (ETT TUKEB, 39258/2019/EKU).

