## Supplementary material for "Socioeconomic inequalities and diabetes complications: An analysis of administrative data from Hungary": Suppl file

### Supplementary files

**Supplementary file Figure S1:** Evolution of aDCSI and the complications around the time of diabetes diagnosis

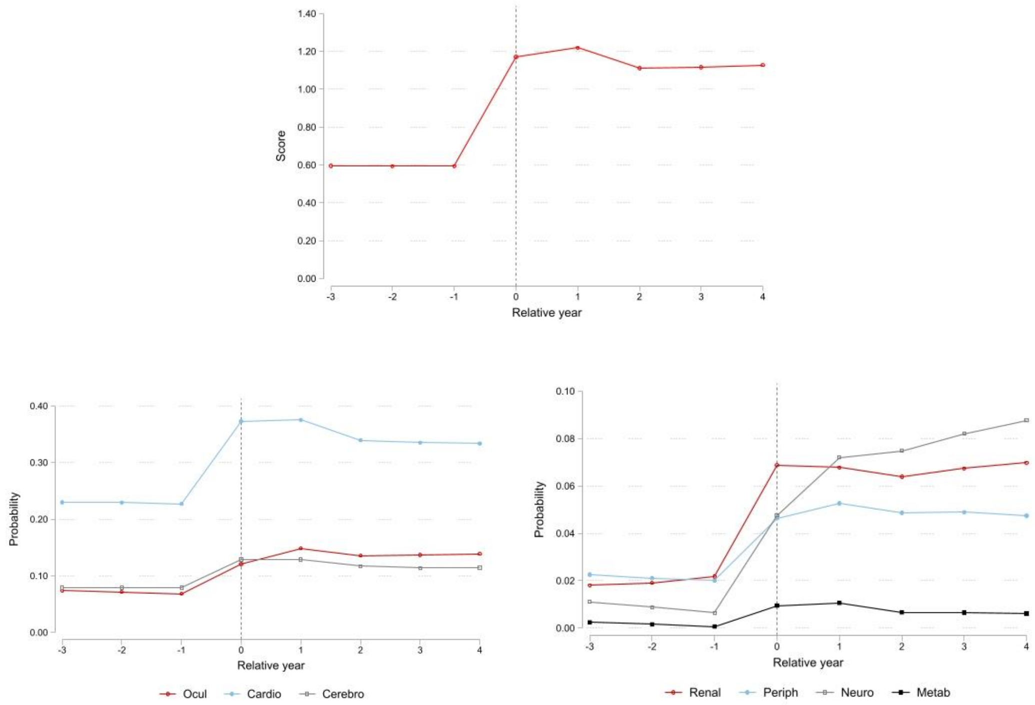


aDCSI score and the probability of diagnosed complications by relative year since diabetes diagnosis. Observation period: 2010-2017.

**Supplementary file Table S1:** Diagnosis codes defining diabetes complications

| Complication type | ICD-10 codes (Hungary) | aDCSI |
|---|---|---|
| Ocular | H350, H358, H353, H356, E103, E113 E123, E133, E143, H280, H360 | 1 |
| | H352, H330, H540, H544, H431 | 2 |
| Renal | E102, E112, E122, E132, E142, N083 N04, N03, N05 | 1 |
| | T824, Z490, Z992 N18, N17, N19, Z49 | 2 |
| Cardiovascular | I70, I24, I20, I25 | 1 |
| | I21, I23, I46, I48, I22, I50, I71 I110, I130, I490 | 2 |
| Cerebrovascular | G45 | 1 |
| | I61, I63, I64, I69 | 2 |
| Peripheral vascular | E105, E115, E125, E135, E145, I792, I724, I739 | 1 |
| | I743, A480, L984 I7024 R02, L97 | 2 |
| Neurological | E104, E114, E124, E134, E144, G609 G730, H490, H491, H492, G590, M146, M142, G632, G990 | 1 |
| Metabolic | E101, E111, E121, E131, E141, E100 E110, E120, E130, E140 | 2 |

ICD-10 diagnosis codes defining diabetes complications by organ system and severity (aDCSI score 1 or 2), following [30]. Within each organ system, the largest score is taken into account.

**Supplementary file Table S2:** Correlation of the presence of diabetes complications

| | (1) | (2) | (3) | (4) | (5) | (6) | (7) |
|---|---|---|---|---|---|---|---|
| (1) Ocular | 1.000 | | | | | | |
| (2) Renal | 0.077 | 1.000 | | | | | |
| (3) Cardiovascular | 0.103 | 0.223 | 1.000 | | | | |
| (4) Cerebrovascular | 0.054 | 0.085 | 0.228 | 1.000 | | | |
| (5) Peripheral vascular | 0.080 | 0.099 | 0.192 | 0.063 | 1.000 | | |
| (6) Neurological | 0.154 | 0.094 | 0.160 | 0.173 | 0.165 | 1.000 | |
| (7) Metabolic | 0.017 | 0.027 | 0.014 | 0.006 | 0.015 | 0.027 | 1.000 |

Correlation between the presence of any organ-specific complications. Population: diabetic patients in 2016.

**Supplementary file Table S3:** Correlation of territorial-level explanatory variables

| | (1) | (2) | (3) | (4) | (5) | (6) | (7) | (8) | (9) |
|---|---|---|---|---|---|---|---|---|---|
| Settlement-level SES variables | | | | | | | | | |
| (1) Unemployment rate (%) | 1.000 | | | | | | | | |
| (2) Ratio of graduates (%) | -0.598 | 1.000 | | | | | | | |
| (3) Log income per cap. | -0.809 | 0.818 | 1.000 | | | | | | |
| Settlement type | | | | | | | | | |
| (4) Budapest | -0.401 | 0.603 | 0.497 | 1.000 | | | | | |
| (5) County seat | -0.089 | 0.289 | 0.239 | -0.204 | 1.000 | | | | |
| (6) Other | 0.387 | -0.705 | -0.582 | -0.627 | -0.635 | 1.000 | | | |
| Healthcare supply variables | | | | | | | | | |
| (7) Filled PCP ratio (%) | -0.243 | 0.213 | 0.254 | 0.095 | 0.099 | -0.154 | 1.000 | | |
| (8) Outpatient hours per cap. | -0.375 | 0.605 | 0.495 | 0.617 | 0.320 | -0.742 | 0.143 | 1.000 | |
| (9) Hospital beds per 1000 cap. | -0.223 | 0.363 | 0.319 | 0.281 | 0.313 | -0.471 | 0.104 | 0.773 | 1.000 |

Correlation between territorial-level demographic, geographic and healthcare supply variables, weighted by the number of diabetic patients. Year: 2016.
